# A higher polygenic score for peripheral artery disease is associated with younger age at surgery among patients undergoing revascularization

**DOI:** 10.64898/2026.06.25.26356619

**Authors:** Jiaqi Hu, Dana Alameddine, Shreef Said, He Wang, Mingfu Yu, Michael Murray, Andrew T. DeWan, Cassius I. Ochoa Chaar

## Abstract

We examined whether polygenic risk for peripheral artery disease (PAD) is associated with severity among patients undergoing lower extremity revascularization at Yale New Haven Hospital. Patients were classified into European (EUR) and non-European (non-EUR) ancestry groups. Associations between the 19-variant polygenic score (PGS) and nine severity indicators were evaluated using linear and Cox regression models stratified by ancestry, followed by meta-analysis. Significant findings (p < 0.05) were assessed for replication in the UK Biobank (UKB). After quality control, 68 EUR and 59 non-EUR patients were included. In EUR patients, higher PGS was associated with increased risk for stroke (HR = 2.43, 95% CI 1.06–5.57). Meta-analysis revealed a significant association between higher PGS and younger age at surgery (β = –2.90, SE = 1.28), which was replicated in the UKB (β = –0.58, SE = 0.15). These results suggest genetic risk contributes to PAD severity.

## Introduction

Peripheral artery disease (PAD) is a debilitating disease affecting over 200 million individuals worldwide[Fowkes et al., 2017]. Patients that present with lifestyle limiting claudication or chronic limb-threatening ischemia (CLTI) require revascularization to improve lower extremity perfusion and represent a subgroup with more advanced disease. Early identification and precise risk stratification are critical to improve the care of patients with PAD[Vartanian and Conte, 2015]. Disease severity and outcomes of revascularization within this subgroup vary due to several key factors, including patient age, metabolic risk factors and their control, lifestyle choices such as smoking, as well as genetic factors that have not been well characterized.

Specific risk factors such as diabetes have shown consistently to be associated with adverse outcomes of revascularization[Singh et al., 2014; Ochoa Chaar et al., 2025]. In addition, patients with premature PAD undergoing revascularization at ≤ 50 years of age experience significantly higher amputation rates, despite aggressive medical and surgical management, compared to older surgical cohorts[Chaar et al., 2012; Kim et al., 2021, 2022]. PAD patients also generally have an elevated risk of major adverse cardiovascular events (MACEs) and major adverse limb events (MALEs)[Hess et al., 2017], although the prevalence and impact of these events differ across individual groups[Hur et al., 2012]. Understanding the genetic mechanisms underlying these factors’ relationship to severity and developing targeted preventive strategies are essential steps toward improving clinical outcomes and reducing the overall burden of PAD.

Emerging evidence suggests a significant genetic component in both the development and progression of PAD[Khaleghi et al., 2014; Matsukura et al., 2015; Klarin et al., 2019; van Zuydam et al., 2021]. A recent genome-wide association study (GWAS), combining data from the Million Veteran Program (MVP) and the UK Biobank (UKB), identified and validated 19 independent single nucleotide polymorphisms (SNPs) associated with the diagnosis of PAD[Klarin et al., 2019]. Moreover, a polygenic score (PGS) developed using the MVP GWAS data and applied to UKB participants demonstrated significant associations with increased PAD risk[Wang et al., 2022] and a higher likelihood of requiring surgical intervention[Hu et al., 2025].

Although at least 19 genetic variants have been identified to be associated with PAD, there is a substantial gap in our understanding of how the cumulative genetic risk from these variants affects disease severity. On the other hand, it is unknown whether the genetic makeup of surgical patients with PAD undergoing revascularization can be leveraged to predict their outcomes or response to various medical or surgical therapies. Specifically, the associations between this aggregated genetic risk and critical factors such as timing and severity of presentation as well as outcomes of reintervention and major amputation, have not been thoroughly investigated. Addressing this significant research gap is essential for enhancing clinical decision making and improving personalized management strategies for PAD patients.

This study explores the associations between a 19-variant PGS and PAD severity, as indicated by revascularization at a premature age, as well as the outcome of revascularization. Our hypothesis is that patients with higher PGS are more likely to have revascularization at a premature age and develop worse outcomes.

## Methods

### Study subjects

Study data were collected and managed using REDCap electronic data capture tools hosted at Yale University[Harris et al., 2009, 2019]. The electronic medical records of patients undergoing lower extremity revascularization (LER) at Yale New Haven Hospital were reviewed between 2011-2020. Only patients who expired and had available pathological specimen from amputation, surgical resection, or biopsies were included. The research protocol was approved by the institutional review board.

The demographics of the patients including age at initial revascularization, sex, and race were recorded. Patient comorbidities included diabetes, hypertension, chronic renal insufficiency defined as baseline creatinine > 1.5 mg/dL, end-stage renal disease (ESRD), coronary artery disease (CAD), congestive heart failure (CHF), stroke, and cancer. Baseline medications including the use of aspirin, antiplatelets, anticoagulants, and statins were captured. The indications for surgery were divided into claudication and chronic limb-threatening ischemia (CLTI). Surgical strategy was divided into open or endovascular. Hybrid surgeries, comprising both an open and an endovascular component, were grouped with the open surgical revascularization group.

### Surgical Outcomes

Outcomes were divided into short-term within the 30-day period after revascularization and long-term (after 30 days). Major amputation (at or above the ankle), reintervention defined by repeat ipsilateral revascularization, as well as myocardial infarction (MI) and stroke were examined. The composite outcomes of MALE defined as major amputation or reintervention and MACE defined as MI or stroke were assessed. The outcome of comprehensive vascular events (CVE) comprised both a combined outcome of MALE or MACE. The outcomes were assessed in the long-term follow up. For long-term follow up, the time to adverse event was analyzed including the time to death.

### Pathological Specimen

In patients who underwent major amputation, the limb specimen preserved in the pathology department were used as primary source for DNA isolation. Other pathology specimen were used if patients did not undergo major amputation. The sources included minor amputation, surgical resection specimen, as well as gastrointestinal and genitourinary biopsies. For each specimen, 20 sections of 10 micrometer thickness (1 curl) were obtained from formaldehyde fixed paraffin embedded (FFPE) block of tissue.

### DNA Extraction and Genotyping

FFPE DNA was isolated from 1-3 curls using Beckman Coulter Formapure Total XL kit as per the manufacturers protocol. The isolated material was quantified using Thermo-Fisher Qubit fluorometric quantification. Illumina FFPE Restoration was then performed according to the manufacturer’s protocol and the resulting repaired material was used as input for Illumina Infinium Global Diversity Array (GDA) processing. The arrays were scanned on an Illumina iScan, the scan data was analyzed using Genome Studio. A total number of 1,914,935 variants were genotyped for four batches.

### Genotype quality control

Genotype data from the four batches were merged using PLINK 1.9[Purcell et al., 2007]. Subsequently, we excluded indels (N=44,172), non-autosomal SNPs (N=74,920), SNPs with ambiguous allele designations (N=134,191), and duplicated SNPs (N=101,596). This filtering yielded a dataset with 1,560,056 SNPs. Additionally, identity-by-descent analysis identified two duplicate pairs; from each pair, we removed the individual with the higher genotype missing rate. Furthermore, one participant with a genotype missing rate exceeding 10% was also excluded. Further quality control (QC) was performed on the remaining 128 subjects by removing SNPs with multiple alleles (N=0) and missing rates greater than 5% (N=137,562).

### Inference of genetic ancestry

Post-QC genotype data were merged with data from 503 EUR-ancestry subjects obtained from the 1000 Genomes Project[1000 Genomes Project Consortium et al., 2015]. The merged dataset was pruned for principal component analysis (PCA). The K-nearest neighbor (KNN) algorithm was then applied to the top 10 PCs to identify outliers relative to the 1000 Genomes EUR reference set[Privé et al., 2020]. Subjects identified as outliers were designated as ‘non-EUR’, while the remaining subjects were classified as ‘EUR’ (Supplemental Figure I).

For each ancestry group, additional QC measures were implemented. Subjects with heterozygosity rates outside the range defined by the mean ± 3 standard deviations were excluded (N=0 for EUR and N=1 for non-EUR). In addition, SNPs with Hardy-Weinberg equilibrium p-values below 5×10^-8^ (N=8,329 for EUR and N=8,041 for non-EUR) or with minor allele frequency (MAF) below 0.001 (N=376,653 for EUR and N=615,118 for non-EUR) were also removed. After applying these criteria, the dataset included 68 EUR subjects with 799,047 SNPs and 59 non-EUR subjects with 1,037,827 SNPs, which were subsequently prepared for imputation.

### Genotype imputation

Imputation was performed using the TOPMed Imputation Server. We imputed the genotypes for 59 non-EUR and 68 EUR subjects using the r3 version of the TOPMed reference panel[Taliun et al., 2019]. Imputation quality was rigorously evaluated using the R^2^ metric, and SNPs with an R^2^ value below 0.3 were excluded from subsequent analyses, ensuring that only high-quality imputed genotypes were incorporated into further analyses.

### UK Biobank

We carried forward to replication our findings that were significant (p-value < 0.05) in any models in EUR-specific analysis and meta-analysis using data from the UKB, a prospective cohort comprising genotype and phenotype information from 502,618 participants recruited from 22 assessment centers across the United Kingdom between 2006 and 2010[Sudlow et al., 2015]. Our analysis focused on unrelated EUR British subjects who underwent surgical intervention for PAD. Subjects were identified based on relevant Office of Population Censuses and Surveys Classification of Interventions and Procedures-4 (OPCS-4) surgical codes[Klarin et al., 2019], and only those with a recorded date of surgery were included in the study. In total, 1,767 subjects met these criteria and were selected for validation. Additional details regarding the specific OPCS-4 codes are provided in Supplemental Table I. In the discovery dataset, the PGS was significantly associated with age at surgery and long-term stroke. Corresponding variables in the UK Biobank were defined using International Classification of Diseases (ICD-9 and ICD-10) codes and Field IDs, as detailed in Supplemental Table I. The diagnosis of stroke was determined using a combination of self-reported data and linked health records, with the corresponding dates of diagnosis extracted from available records. As the UKB does not provide exact dates for self-reported diagnoses, these were imputed as the first day of the recorded month in this study. Notably, individuals who experienced either outcome within one month following surgery or had missing date at diagnosis were excluded from the respective outcome analyses.

### PGS development

The PGS for each subject was computed as the weighted sum of the number of minor alleles across 19 previously identified variants scaled by the total number of SNPs using PLINK 1.9. The weights, corresponding to beta coefficients, were derived from the MVP EUR GWAS summary statistics for PAD, which was based on 24,009 PAD cases and 150,984 non-PAD controls[Klarin et al., 2019]. This summary statistic was obtained from the Database of Genotype and Phenotype (dbGaP; accession number 32518). To facilitate subsequent analyses, the final PGS was standardized to a mean of zero and a standard deviation of one across all study subjects.

### Statistical analysis

Ancestry-specific analyses were conducted to calculate genetic PCs as covariates. Specifically, we began by pruning variants for linkage disequilibrium using PLINK-1.90. A sliding window of 50 variants was applied with a pairwise r^2^ threshold of 0.2, and the window was shifted by 5 variants after each iteration, yielding 219,499 independent variants for EUR and 365,421 for non-EUR. PCA was then performed on this LD-pruned dataset using the smartPCA program in EIGENSOFT-7.2.1[Patterson et al., 2006].

Variation in the constructed PGS across genotype batches and predicted ancestry groups were assessed using ANOVA and logistic regression, respectively.

The associations between the PGS and outcome variables were assessed via Cox proportional hazards models stratified by genetically predicted ancestry. Similarly, associations between the PGS and age at surgery were evaluated via linear regression. Two models with varying levels of covariate adjustment were evaluated: (1) a crude model with no covariates; (2) a demographic model adjusting for age, sex, top four PCs, body mass index (BMI), and smoking. Ancestry-specific association estimates were then combined using a fixed-effect meta-analysis model.

A significance level of 0.05 was used throughout, and all association analyses were conducted using R version 4.2. To be considered replicated, results needed to be significant (p<0.05), and effect estimates in the same direction.

## Results

### Baseline characteristics (Table I)

After QC, the final sample comprised 68 EUR and 59 non-EUR patients, with demographic and clinical characteristics detailed in Table I. The mean age of the population was 66 with no significant difference between the 2 groups. There was a trend towards EUR patients being more likely to be males compared to non-EUR but that did not reach statistical significance (76% vs

**Table I.**
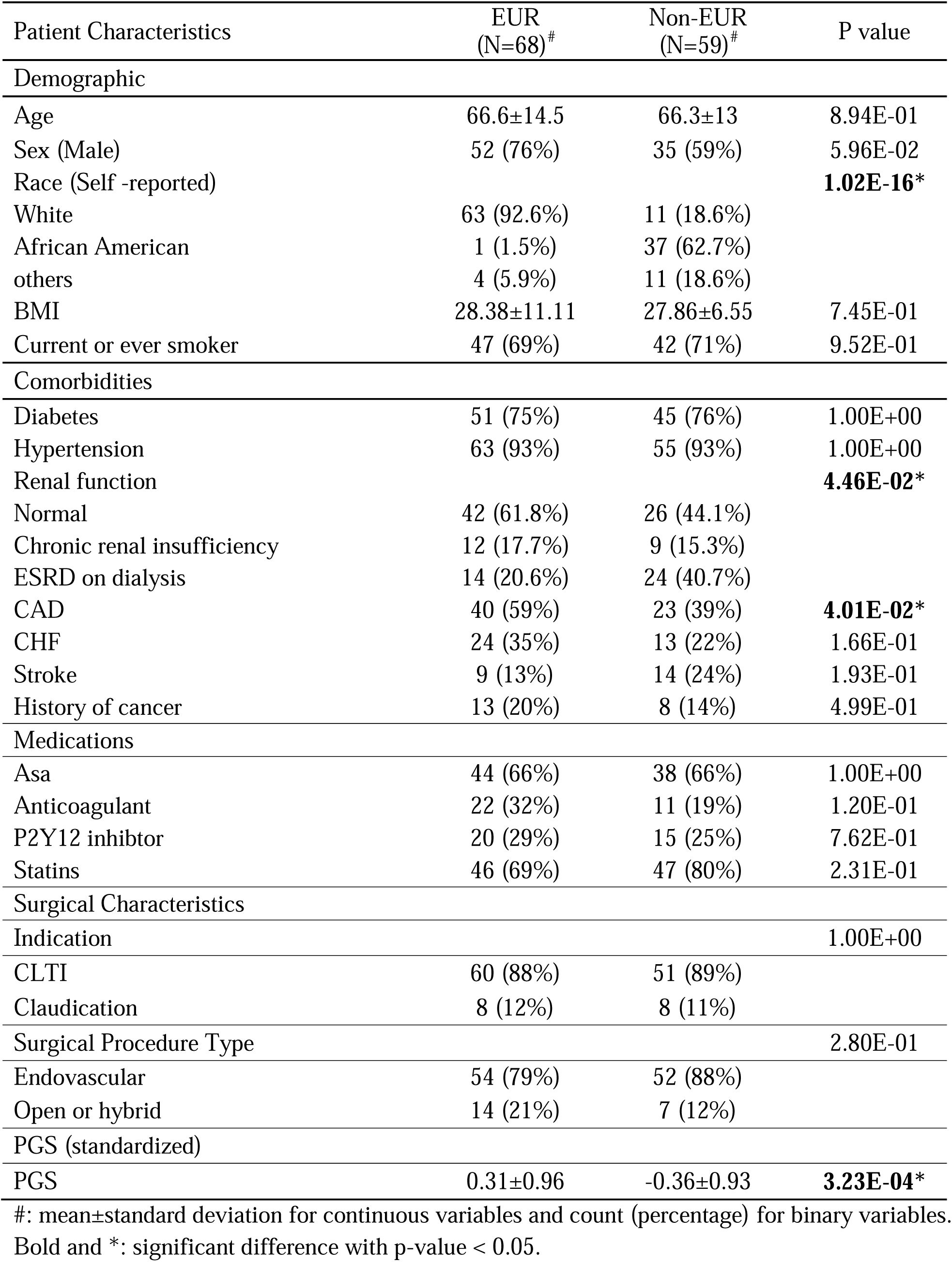
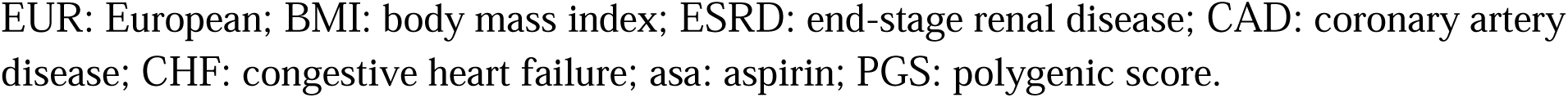
Demographic characteristics and comorbidities.

| Patient Characteristics | EUR<br>(N=68)<sup>#</sup> | Non-EUR<br>(N=59)<sup>#</sup> | P value |
| --- | --- | --- | --- |
| **Demographic** |  |  |  |
| Age | 66.6±14.5 | 66.3±13 | 8.94E-01 |
| Sex (Male) | 52 (76%) | 35 (59%) | 5.96E-02 |
| Race (Self -reported) |  |  | **1.02E-16\*** |
| White | 63 (92.6%) | 11 (18.6%) |  |
| African American | 1 (1.5%) | 37 (62.7%) |  |
| others | 4 (5.9%) | 11 (18.6%) |  |
| BMI | 28.38±11.11 | 27.86±6.55 | 7.45E-01 |
| Current or ever smoker | 47 (69%) | 42 (71%) | 9.52E-01 |
| **Comorbidities** |  |  |  |
| Diabetes | 51 (75%) | 45 (76%) | 1.00E+00 |
| Hypertension | 63 (93%) | 55 (93%) | 1.00E+00 |
| Renal function |  |  | **4.46E-02\*** |
| Normal | 42 (61.8%) | 26 (44.1%) |  |
| Chronic renal insufficiency | 12 (17.7%) | 9 (15.3%) |  |
| ESRD on dialysis | 14 (20.6%) | 24 (40.7%) |  |
| CAD | 40 (59%) | 23 (39%) | **4.01E-02\*** |
| CHF | 24 (35%) | 13 (22%) | 1.66E-01 |
| Stroke | 9 (13%) | 14 (24%) | 1.93E-01 |
| History of cancer | 13 (20%) | 8 (14%) | 4.99E-01 |
| **Medications** |  |  |  |
| Asa | 44 (66%) | 38 (66%) | 1.00E+00 |
| Anticoagulant | 22 (32%) | 11 (19%) | 1.20E-01 |
| P2Y12 inhibitor | 20 (29%) | 15 (25%) | 7.62E-01 |
| Statins | 46 (69%) | 47 (80%) | 2.31E-01 |
| **Surgical Characteristics** |  |  |  |
| Indication |  |  | 1.00E+00 |
| CLTI | 60 (88%) | 51 (89%) |  |
| Claudication | 8 (12%) | 8 (11%) |  |
| Surgical Procedure Type |  |  | 2.80E-01 |
| Endovascular | 54 (79%) | 52 (88%) |  |
| Open or hybrid | 14 (21%) | 7 (12%) |  |
| **PGS (standardized)** |  |  |  |
| PGS | 0.31±0.96 | -0.36±0.93 | **3.23E-04\*** |

<sup>#</sup>: mean±standard deviation for continuous variables and count (percentage) for binary variables.
Bold and \*: significant difference with p-value < 0.05.
EUR: European; BMI: body mass index; ESRD: end-stage renal disease; CAD: coronary artery disease; CHF: congestive heart failure; asa: aspirin; PGS: polygenic score.

59%, P=5.96E-02). There were significant differences in the self-reported race between the 2 groups with the majority of EUR being White (92.6%) while the non-EUR being African American (62.7%) (P<1.00E-03). Patients with EUR ancestry were significantly more likely to have CAD (59% vs 39%, P=4.01E-02) but less likely to have ESRD requiring dialysis (20.6% vs 40.7%, P=4.46E-02) compared to patient with non-EUR ancestry. The majority of patients had diabetes (75%) and hypertension (93%) with no significant difference between the 2 groups.

There was no difference in the medication prescribed for atherosclerosis prior to lower extremity revascularization with 66% on aspirin and 69-80% on statin therapy. Most patients were treated for CLTI (88%) with initial endovascular revascularization (79%-88%) with no statistically significant difference between the 2 groups. The PGS was significantly higher in patients with EUR compared to patients with non-EUR ancestry (0.31 vs -0.36, P=3.23E-04).

### Outcomes (Table II)

**Table II.**
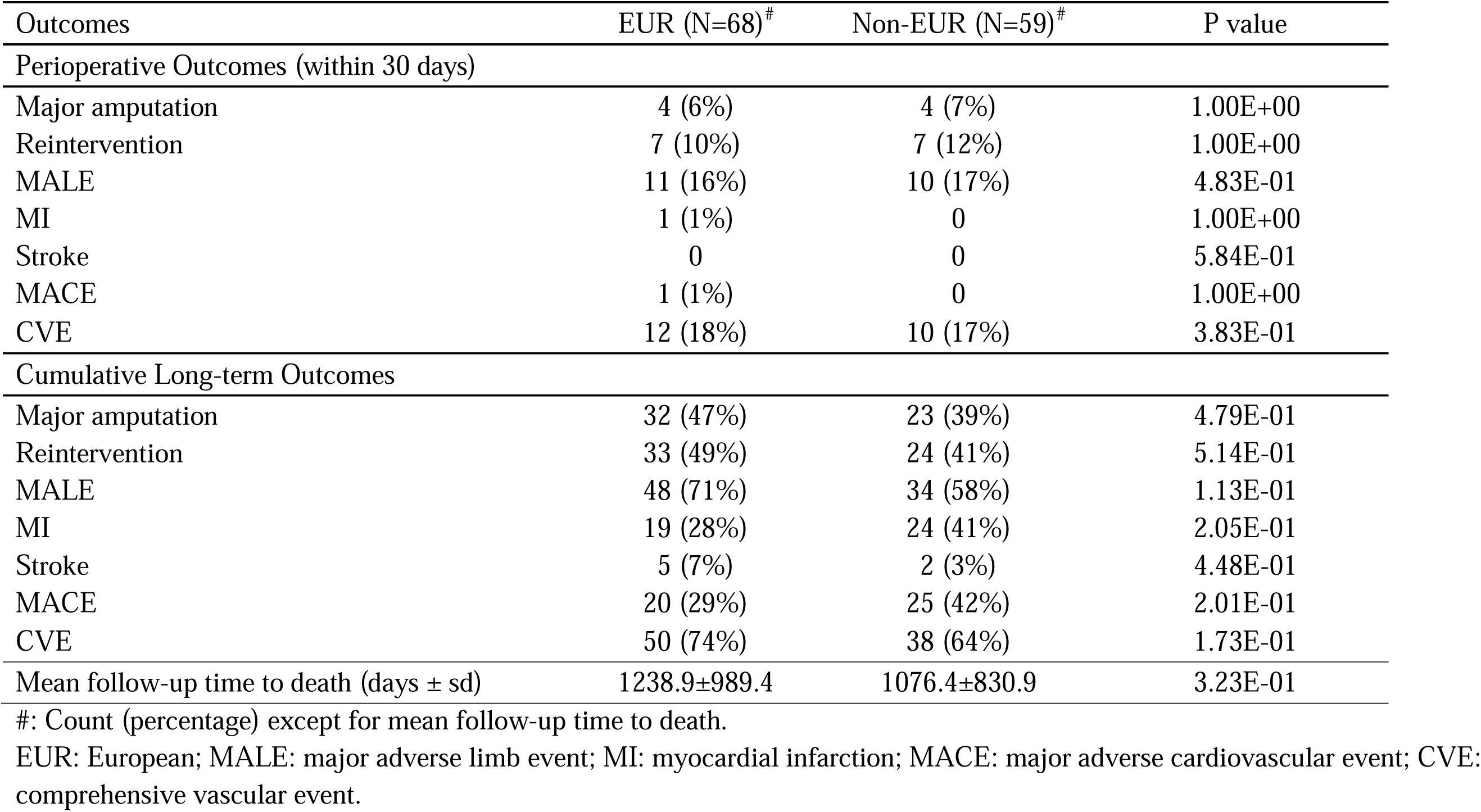
Outcomes.

| Outcomes | EUR (N=68)<sup>#</sup> | Non-EUR (N=59)<sup>#</sup> | P value |
| --- | --- | --- | --- |
| Perioperative Outcomes (within 30 days) |  |  |  |
| Major amputation | 4 (6%) | 4 (7%) | 1.00E+00 |
| Reintervention | 7 (10%) | 7 (12%) | 1.00E+00 |
| MALE | 11 (16%) | 10 (17%) | 4.83E-01 |
| MI | 1 (1%) | 0 | 1.00E+00 |
| Stroke | 0 | 0 | 5.84E-01 |
| MACE | 1 (1%) | 0 | 1.00E+00 |
| CVE | 12 (18%) | 10 (17%) | 3.83E-01 |
| Cumulative Long-term Outcomes |  |  |  |
| Major amputation | 32 (47%) | 23 (39%) | 4.79E-01 |
| Reintervention | 33 (49%) | 24 (41%) | 5.14E-01 |
| MALE | 48 (71%) | 34 (58%) | 1.13E-01 |
| MI | 19 (28%) | 24 (41%) | 2.05E-01 |
| Stroke | 5 (7%) | 2 (3%) | 4.48E-01 |
| MACE | 20 (29%) | 25 (42%) | 2.01E-01 |
| CVE | 50 (74%) | 38 (64%) | 1.73E-01 |
| Mean follow-up time to death (days ± sd) | 1238.9±989.4 | 1076.4±830.9 | 3.23E-01 |

<sup>#</sup>: Count (percentage) except for mean follow-up time to death.
EUR: European; MALE: major adverse limb event; MI: myocardial infarction; MACE: major adverse cardiovascular event; CVE: comprehensive vascular event.

In the perioperative period, there was no difference between the 2 groups in terms of complications. The major amputation at 30 days was 6-7% with 10-12% reintervention rate. Only one patient had a myocardial infarction in the EUR group with no strokes recorded. After 3 years of follow up, the cumulative rates of major amputations (39% vs 47%, P=4.79E-01), reinterventions (41% vs 49%, P=5.14E-01), MI (28% vs 41%, P=2.05E-02), and stroke (7% vs 3%, P=4.48E-01) were not significantly different between the 2 groups. The combined outcomes of MALE, MACE, and CVE were not significantly different neither.

### PGS development

Following genotype imputation, all 19 candidate variants in the EUR subjects met the R^2^ threshold of 0.3; however, one single nucleotide polymorphism (SNP), rs6025, had an R^2^ of 0.014 and was excluded from the PGS calculations for non-EUR.

### Association of PGS with outcomes

The ANOVA showed no significant differences in the PGS across the four genotype batches (P= 5.36E-01); however, the PGS was significantly higher in the predicted EUR group compared to the non-EUR group (P=3.23E-04). In light of this finding, subsequent analyses were stratified by genetic ancestry. Given the heterogeneity of ancestries among non-EUR group (Table I) and the insufficient sample sizes to further identify more refined ancestry groups, the following analysis focused on results for EUR group and associated meta-analysis. One association with PGS reached significance in the EUR-specific analyses: the positive association with long-term stroke in the crude model with a HR of 2.43 (95% CI: 1.06-5.57, P=3.66E-02) (Table III). The model failed to converge after adding the covariates. Other associations remained non-significant (Supplemental Table II). Meta-analyzing EUR and non-EUR generated another significant association, where an increased PGS was associated with a decreased age at surgery in the demographic model (β=-2.9, SE = 1.28, P=2.31E-02). No other significant associations were found in meta-analysis (Supplemental Table II).

**Table III.** Association results.

| Dependent variable | Model | EUR |  | Meta-analysis |  | Replication |  |
| --- | --- | --- | --- | --- | --- | --- | --- |
|  |  | Effect size<sup>#</sup> | P-value | Effect size | P-value | Effect size | P-value |
| Stroke | Crude | 2.43 (1.06-5.57) | 3.66E-02* | 1.71 (0.82-3.55) | 1.50E-01 | 1.08 (0.13-9.02) | 7.98E-01 |
|  | Demographic | Failed to converge |  |  |  | 1.05 (0.13-8.28) | 4.95E-01 |
| Age at surgery | Crude | -2.72±1.76 | 1.27E-01 | -2.28±1.22 | 6.29E-02 | -0.40±0.21 | 5.02E-02 |
|  | **Demographic** | **-3.09±1.89** | **1.08E-01** | **-2.9±1.28** | **2.31E-02\*** | **-0.58±0.15** | **1.01E-04\*** |

EUR: European ancestry.
<sup>#</sup>: HR (95% CI) for stroke and beta±standard error for age at surgery.
\*: statistical significance ( $p < 0.05$ ).
Bold: Replicated finding.

### Replication in the UK Biobank

To validate the significant associations observed in the EUR-specific analysis and meta-analysis—including those with stroke and age at surgery—we repeated the analysis in a cohort of 1,767 unrelated EUR PAD patients with documented surgical procedure codes. Cox proportional hazards models were fitted for 97 patients of stroke diagnosed at or after 30 days of surgery and 1,519 controls. Linear regressions were fitted for age at surgery. The same two sets of covariates were adjusted. Significant associations were replicated for age at surgery in the demographic model (β = –0.58, SE = 0.15, P=1.01E-04). Other associations did not reach statistical significance (P>0.05; Table III).

## Discussion

In this study, we examined the association between aggregated genetic risk, represented by a PGS, and indicators of disease severity as well as outcomes of revascularization in PAD patients who underwent LER. Our findings demonstrate a robust and replicable association between higher genetic risk and earlier age at surgery, supporting the hypothesis that increased genetic susceptibility contributes to earlier presentation of more severe disease and likely faster progression requiring LER. Although our discovery analysis identified potential PGS associations for stroke occurring after 30 days of follow-up, these findings were not replicated in an independent dataset with a larger sample size, highlighting the need for additional research to confirm and clarify the role of genetic risk in these outcomes following PAD surgery.

Our findings align with existing evidence that genetic predisposition contributes to earlier onset and increased severity of vascular diseases. Mars et al (2020) reported a higher frequency of elevated genetic risk among patients with early-onset coronary artery disease and atrial fibrillation. This pattern has also been observed in type 2 diabetes[Mars et al., 2020] and various cancers [Hu et al., 2024]. Besides, prior studies have consistently demonstrated that genetic risk factors can accelerate PAD progression and severity[Khaleghi et al., 2014; Klarin et al., 2019].

Collectively, these findings underscore the potential value of incorporating genetic screening into personalized risk assessments, facilitating improved early detection and individualized management strategies for PAD.

Our previous analysis examining the associations between the 19-variant PGS and PAD severity in the UKB found a significant relationship with the requirement for surgical intervention but did not detect a significant association with premature PAD (defined as age of onset less than or equal to 55)[Hu et al., 2025]. The discrepancy between those results and the current findings could be explained by several factors. Firstly, differences in the definition and measurement of age are crucial. The previous analysis evaluated the age at initial PAD diagnosis, while the current study assessed age at the time of surgical intervention among PAD patients who required surgery. These two age-related metrics, though both indicative of PAD severity, inherently capture different clinical and genetic dimensions. Our findings suggest that age at surgery may be more strongly correlated with genetic predisposition to disease severity compared to age at diagnosis. Additionally, the typically asymptomatic nature of PAD may result in delayed diagnoses, potentially biasing associations towards null results by artificially inflating the age at diagnosis. Therefore, age at surgery may represent a more accurate and clinically relevant measure of disease severity. Collectively, our current findings reinforce the importance of considering surgical timing as an informative indicator of PAD severity linked to genetic risk, aligning closely with and extending insights from our previous research.

The associations observed between our PAD PGS and stroke, which was identified in our discovery analysis, despite not being replicated with the same directions in the UKB, require careful interpretation. The failure to replicate could be attributed to several factors. First, our initial positive result might represent a false-positive finding due to a smaller sample size and limited statistical power, increasing the risk of type I error. Second, differences in definitions for these outcomes and population characteristics, including genetic backgrounds, prevalence of clinical risk factors, variations in clinical management, or environmental exposures, could have influenced the reproducibility of genetic associations across datasets. These factors underscore the importance of replication studies and highlight the necessity for further large-scale analyses. Such research is essential for accurately delineating the relationship between genetic predisposition to PAD and outcomes after surgical intervention, ultimately facilitating more precise risk stratification and targeted preventative strategies.

This study has several limitations that should be acknowledged. First, the sample size is limited and we may lack the statistical power in the analyses. Due to that, we adopted a relaxed significance threshold (P-value < 0.05) without correction for multiple comparisons, increasing the risk of type I error. Nevertheless, the replication analyses conducted using the larger UKB cohort help mitigate this concern by providing additional statistical power and validation. Second, the genetic ancestry stratification used in this study categorized participants into EUR and non-EUR groups. Interestingly, not all EUR identified themselves as White and the non-EUR group had significant heterogeneity that interfered with a genetic analysis due heterogeneity and reduced our sample size significantly. Third, potential selection bias may have been introduced during sample collection, as patients who underwent amputation were more likely to have tissue specimens available. Thus the high percentage of MACE and MALE in all patients after only 3 years of follow up. Fourth, the number of MACE events may have been underestimated due to the potential for deaths occurring outside the hospital setting. Fifth, the self-reported dates of stroke diagnosis in the UKB may be imprecise, potentially introducing bias into the analysis. Nonetheless, because the association with stroke was not replicated in the UKB, this limitation is unlikely to substantially affect our conclusions. Lastly, our research is exploratory and hypothesis-generating by design; therefore, additional studies with larger sample sizes, precise ancestry definitions, and rigorous statistical methods are required to confirm and further investigate the genetic associations and hypotheses proposed in our analyses.

## Conclusion

We identified and replicated a significant association between increased genetic risk for PAD and an earlier age at surgery, highlighting the potential link between genetic factors and disease severity. Our findings offer valuable insights that could enhance clinical decision-making and inform personalized treatment strategies for PAD patients requiring surgical interventions.

## Data Availability

The datasets used and/or analyzed during the current study are available from the corresponding author on reasonable request.

## List of abbreviations

PAD: peripheral artery disease
CLTI: chronic limb-threatening ischemia
MACE: major adverse cardiovascular event
MALE: major adverse limb event
CVE: comprehensive vascular event
GWAS: genome-wide association study
MVP: Million Veteran Program
UKB: UK Biobank
SNP: single nucleotide polymorphism
PGS: polygenic score
LER: lower extremity revascularization
ESRD: end-stage renal disease
CAD: coronary artery disease
CHF: congestive heart failure
MI: myocardial infarction
GDA: Global Diversity Array
QC: quality control
PCA: principal component analysis
KNN: K-nearest neighbor
EUR: European
MAF: minor allele frequency
SD: standard deviation

## Declarations

### Ethics approval and consent to participate

This research was conducted using the UK Biobank Resource (application number 32285). The UK Biobank study was conducted under generic approval from the National Health Services’ National Research Ethics Service. The present analyses were conducted in accordance with the Declaration of Helsinki and approved by the Human Investigations Committee at Yale University (2000026836 for UK Biobank). The IRB provided waiver of signed consent.

### Consent for publication

Not applicable.

### Authors’ contributions

JH, DA, CIOC, and AD contributed to the study conception and design. Data requests and analysis of the discovery datasets were performed by JH, DA, CIOC, and AD, SS, HW, MY and CIOC contributed to the data collection and processing. The first draft of the manuscript was written by JH, CIOC, and AD, and all authors commented on the manuscript. All authors read and approved the final manuscript.

## Acknowledgements

The authors would like to thank the researchers and participants of the United Kingdom Biobank. All data was accessed as part of project 32285 from the United Kingdom Biobank. We thank the Yale Center for Research Computing for the use of the McCleary High Performance Computing cluster.

## Declaration of Generative AI in Scientific Writing

During the preparation of this work the author(s) used ChatGPT in order to improve readability. After using this tool/service, the author(s) reviewed and edited the content as needed and take(s) full responsibility for the content of the publication.

