## Supplementary materials for "A higher polygenic score for peripheral artery disease is associated with younger age at surgery among patients undergoing revascularization"

Supplemental Materials

Supplemental Table I Definition codes in UKB

|  | Field ID | ICD-10 | ICD-9 | OPCS-4 |
| --- | --- | --- | --- | --- |
| PAD patients who received surgery |  |  |  | X09.3, X09.4, X09.5 L21.6, L51.3, L51.6, L51.8, L52.1, L52.2, L54.1, L54.4, L54.8 L59.1, L59.2, L59.3, L59.4, L59.5, L59.6, L59.7, L59.8, L60.1, L60.2,  L63.1, L63.5, L63.9, L66.7 |
| Sex | 31 |  |  |  |
| BMI | 21001 |  |  |  |
| Current smoking | 20116 (2) |  |  |  |
| Ever smoking | 20116 (1) |  |  |  |
| Stroke | 20002 (1583, 1491, 1081, 1086) | I60.0-I60.9, I61.0-I61.9, I62.0, I62.1, I62.9, I63.0-I63.9, I64 | 430.9, 431.9, 434.9, 436.9 |  |

Supplemental Table II Association results

|  |  | EUR | | Meta-analysis | | |
| --- | --- | --- | --- | --- | --- | --- |
| Outcome | Model | HR (95% CI) | P-value | | HR (95% CI) | P-value |
| Major amputation | Crude | 0.86 (0.6-1.23) | 4.08E-01 | | 1.12 (0.84-1.49) | 4.57E-01 |
|  | Demographic | 0.91 (0.62-1.34) | 6.42E-01 | | 1.25 (0.91-1.72) | 1.61E-01 |
| Reintervention | Crude | 1.01 (0.71-1.43) | 9.63E-01 | | 1.10 (0.83-1.44) | 5.14E-01 |
|  | Demographic | 1.13 (0.78-1.64) | 5.21E-01 | | 1.10 (0.81-1.49) | 5.61E-01 |
| MALE | Crude | 0.80 (0.58-1.11) | 1.81E-01 | | 1.01 (0.79-1.29) | 9.32E-01 |
|  | Demographic | 0.92 (0.65-1.30) | 6.20E-01 | | 1.08 (0.82-1.42) | 5.97E-01 |
| MI | Crude | 1.25 (0.82-1.91) | 3.04E-01 | | 0.95 (0.70-1.27) | 7.16E-01 |
|  | Demographic | 0.93 (0.58-1.49) | 7.59E-01 | | 0.81 (0.57-1.15) | 2.37E-01 |
| MACE | Crude | 1.31 (0.87-1.98) | 1.94E-01 | | 0.95 (0.71-1.27) | 7.46E-01 |
|  | Demographic | 1.00 (0.63-1.59) | 9.95E-01 | | 0.84 (0.59-1.19) | 3.24E-01 |
| CVE | Crude | 0.83 (0.61-1.12) | 2.21E-01 | | 0.94 (0.74-1.18) | 5.74E-01 |
|  | Demographic | 0.90 (0.64-1.26) | 5.41E-01 | | 0.97 (0.75-1.26) | 8.20E-01 |
| Time to death | Crude | 0.84 (0.66-1.08) | 1.73E-01 | | 0.88 (0.74-1.05) | 1.62E-01 |
|  | Demographic | 0.84 (0.62-1.12) | 2.33E-01 | | 0.98 (0.80-1.21) | 8.65E-01 |

Supplemental Figure I. Ancestry inference using PCA.

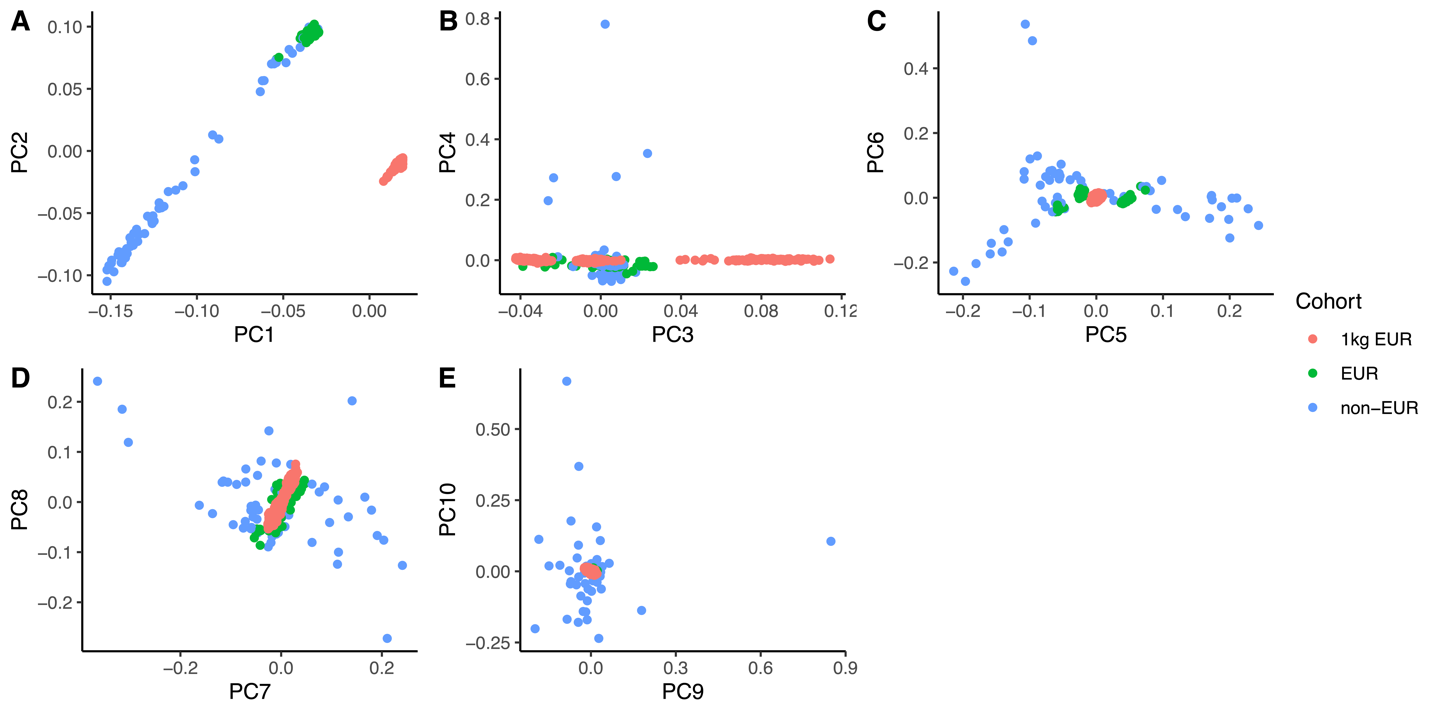
